# Language areas revisited by principal component analysis of electrical cortical stimulation mapping

**DOI:** 10.1101/2025.03.21.25323362

**Authors:** Mayumi Otani, Akihiro Shimotake, Takuro Nakae, Mitsuhiro Sakamoto, Masao Matsuhashi, Takayuki Kikuchi, Kazumichi Yoshida, Takeharu Kunieda, Susumu Miyamoto, Ryosuke Takahashi, Matthew A Lambon-Ralph, Akio Ikeda, Riki Matsumoto

## Abstract

Electrical cortical stimulation (ECS) remains the gold standard for language mapping in neurosurgery; thus, systematic and less invasive language tasks focusing on each language function should be developed. This study aimed to clarify the correlation between anatomy and language function by extracting principal components (PCs) directly related to language function from six tasks. Twelve patients with intractable temporal lobe epilepsy underwent ECS mapping with subdural electrode placement in the language-dominant hemisphere: 313 electrodes (3–48/patient) were confirmed to be located over the language areas by ECS mapping using six language tasks. Three major PCs were delineated; PC1, PC2, and PC3 represented reading, auditory receptive semantic processing, and expressive semantic processing, respectively. Anatomically, PC1 was prominent at the inferior frontal gyrus (IFG, especially pars opercularis), posterior parts of the superior temporal sulcus (STS), middle temporal gyrus (MTG), and inferior temporal gyrus; PC2 at the posterior part of IFG (extended more anteriorly than PC1), posterior superior temporal gyrus (STG), posterior MTG, and anterior ventral temporal cortex (VTC); and PC3 at the broad area including regions of the PC1 and PC2, and supramarginal gyrus (SMG) and VTC. Statistically significant functional differentiation was observed in the posterior language area (between the SMG and posterior STG) and basal temporal language areas (among the anterior, middle, and posterior parts). In conclusion, PC analysis revealed three independent language functions and indicated functional differences within the posterior and basal temporal language areas. Clinically efficient language mapping is expected by task selection, considering PCs and regions.

## 1 Introduction

Human language areas and language networks have traditionally been studied mainly in the context of aphasia caused by stroke and have been formally related to perisylvian language areas (Geschwind, 1965). Classically, Broca’s area (Brodmann areas 44 and 45, called the anterior language area), centered on the posterior part of the inferior frontal gyrus (IFG), was associated with articulation, but is now understood to be linked with grammar and semantics. Wernicke’s area, or the posterior language area together with the surrounding supramarginal gyrus (SMG) and/or angular gyrus, is also thought to be involved in the storage and retrieval of phonological information and has a role in language perception. In the classical language model, which has been the fundamental idea on language network to date, the two language areas are connected by the arcuate fasciculus. In the 1990s, the basal temporal language area (BTLA) as the third language area (Luders et al., 1991) and multiple language networks beyond the perisylvian region itself were elucidated using various methods, such as fine-scale functional localization (Catani et al., 2005; Hickok & Poeppel, 2007), functional magnetic resonance imaging (Price, 2012), explorations of primary progressive aphasia (Gorno-Tempini et al., 2011; Hodges & Patterson, 2007; Lambon Ralph et al., 2001) and electrocorticographic mapping using high gamma activity (Crone et al., 2006; Mitchell et al., 2008). Despite the increasing number of new methods, directly identifying language areas by electrical stimulation testing remains the gold standard.

For language mapping by electrical cortical stimulation, it is important to select tasks that can be completed within the duration of electrical stimulation, are less effortful, and can be performed repeatedly; therefore, simple tasks, such as picture naming, are most often used. Many studies have compared picture naming with other tasks to estimate language function; however, when naming is used alone, attempts have been made to clarify the localization of language function by analyzing and comparing the patients’ paraphasias (Corina et al., 2010; Perrone-Bertolotti et al., 2020). This method often raises concerns due to the potential for substantial subjectivity from both the participants and the evaluators; plus demonstrations that there is no one-to-one mapping between error type and functional locus of damage(Halai et al., 2018). In contrast, this study utilized a unique and powerful opportunity in which patients are assessed across a test battery rather than a single task. Specifically, patients completed a detailed and accurate preoperative evaluation that enables complementary assessment of function by mapping the Japanese language using six different tasks. These tasks were set with attention to semantic processing, taking advantage of the characteristics of the Japanese language, which has ideographic characters (kanji). A previous study on event-related potentials for Japanese reading revealed differences in the reading-related areas for ideographic and phonographic (kana) characters, indicating that employing tasks that utilize both kanji and kana can be instrumental in enhancing our understanding of language functions (Usui et al., 2003). Clinically, multiple tasks also contribute to mapping and evaluating the core language area, and detecting areas to be preserved during epilepsy surgery.

A potential limitation of comprehensive assessment batteries is that the number of tasks becomes too demanding to the patient and clinically impractical to administer. An intermediate approach, therefore, is to comprehensively assess the anatomo-functional relationship of the language system with fewer examinations combined with more advanced statistical approaches. (Halai et al., 2022)Accordingly, the objectives of the present study were the following: 1) to extract principal components (PCs) directly related to language function from the six tasks for more efficient and effective task selection; and 2) to clarify the correlation between anatomy and language function that may be equivalent to each PC. Specifically, we conducted principal component analysis (PCA), visualization of PC scores, and comparison of PC scores across domains.

## 2 Materials and methods

### 2.1 Participants

We recruited 12 patients with intractable partial epilepsy (epilepsy due to brain tumor, n=4) who underwent subdural electrode implantation in the language dominant hemisphere (left [all patients except patient 9] or right [patient 9]) for presurgical evaluation. Patients with broad electrode coverage in the language-related area, that is, patients with multiple electrodes placed in at least two of the three locations of the frontal lobe (which includes the anterior language area), lateral temporal and parietal lobes (posterior language area), and basal temporal lobe (basal temporal language area), were included. The subdural electrodes were made of platinum with an interelectrode distance and recording diameter of 1 cm and 2.3 mm (Ad-Tech Medical Instrument Corporation, Oak Creek, WI, USA), respectively. The patients’ demographics are summarized in Table 1. This study was approved by the ethics committee of our institute (approval number, C533), and informed consent was obtained from all the patients.

**Table 1.** The demographics and clinical information of the patients. Ages are presented in non-overlapping ranges (minimum 5-year intervals) to ensure patient confidentiality.

| Patient | 1 | 2 | 3 | 4 | 5 | 6 |
| --- | --- | --- | --- | --- | --- | --- |
| Age(range), gender, handedness | 21-25, M, R | 25-30, M, R&L | 16-20, F, R | 36-40, F, R | 51-55, M, R | 51-55, M, R |
| WAIS-III (VIQ, PIQ, TIQ) | 70, 78, 69 | 72, 78, 72 | 67, 76, 69 | 84, 97, 89 | 105, 99, 103 | 73, 97, 83 |
| WMS-R(Verbal, Visual, General, Attention, Delayed recall) | 99,64,87, 91, 82 | 99, 92, 97, 87, 83 | 51, <50, <50, 81, 56 | 75, 111, 83, 62, 53 | 71, 117, 84, 109, 72 | 80, 101, 85, 101, 91 |
| WAB | 95.6 | 96 | 97.2 | 98.5 | 98 | 89.6 |
| WADA test (Language) | Left | Bilateral | Left | Left | Left | Left |
| Age(range) of seizure onset | 16-20 | 6-10 | 10-15 | 26-30 | 51-55 | 41-45 |
| Ictal ECoG onset | frontal&temporal lobe | Lateral&mesial temporal lobe | Mesial temporal lobe | Mesial temporal lobe | None | Lateral temporal lobe |
| Pathology | FCD type 1A | FCD, Hippoampal sclerosis | FCD | FCD | Diffuse astrocytoma | Arteriovenous malformation gliosis |

| Patient | 7 | 8 | 9 | 10 | 11 | 12 |
| --- | --- | --- | --- | --- | --- | --- |
| Age(range), gender, handedness | 61-65, M, R | 26-30, F, R | 36-40, M, L | 21-25, M, R | 41-45, M, R | 31-35, M, R |
| WAIS-III (VIQ, PIQ, TIQ) | 109, 115, 112 | 62, 80, 67 | 93, 105, 98 | 86, 79, 81 | 96, 84, 90 | 82, 86, 82 |
| WMS-R(Verbal, Visual, General, Attention, Delayed recall) | 71, 79, 70, 90, 58 | 64, 94, 68, 79, 79 | 74, 94, 77, 110, 96 | 55, 79, 53, 90, 54 | 73, 85, 73, 103, 76 | 75, 89, 75, 92, 81 |
| WAB | 96.9 | 95.8 | 99 | 97.4 | 99.9 | 99.2 |
| WADA test (Language) | Left | Left | Right | Left | Bilateral (L>R) | Left |
| Age(range) of seizure onset | 26-30 | 10-15 | 6-10 | 10-15 | 26-30 | 20-25 |
| Ictal ECoG onset | lateral&mesial temporal lobe | mesial temporal lobe | mesial temporal lobe | lateral&mesial temporal lobe | mesial temporal lobe | mesial temporal lobe |
| Pathology | Oligoastrocytoma | None | FCD, HS | FCD type | FCD, HS | FCD |

### 2.2 Functional cortical mapping

#### 2.2.1 Apparatus

High-frequency electrical cortical stimulation was performed using subdural electrodes. Repetitive, square-wave electric currents of alternating polarity with a pulse width and frequency of 0.3 ms and 50 Hz, respectively, were delivered through a pair of electrodes for 1–5 s (Electrical stimulator MS-120B; Nihon Kohden, Tokyo, Japan). Details of the methodology for cortical stimulation and the subsequent cortical mapping have been described elsewhere (Matsumoto et al., 2011). Language electrode refers to the electrode where the cortex (directly below) is assessed as the language area. The assessment criteria are described below.

#### 2.2.2 Assessment criteria

For mapping each area of the brain, after confirming the absence of positive (e.g., tonic contraction) and negative (e.g., impairment of rapid alternating movements) tongue motor responses, a series of semantic and language assessments were tested concurrently with a 4–5 s period of electrical stimulation at 10–15 mA. Responses to the assessment battery shown below were rated from 0–2 points based on participant errors during stimulation (Matoba et al., 2024; Shimotake et al., 2015; Usui et al., 2003). Two points were awarded for no response (arrest) and one point for a delay in response (slowing) or an incorrect verbal reply (error). We considered the impaired behaviors as significant when the findings were reproducible in the absence of afterdischarges. When frequent after-discharges occurred, we decreased stimulation intensity to 8 or 9 mA. All sessions were video-recorded, and electrocorticography was recorded simultaneously.

##### Paragraph reading

The patients were asked to read aloud a part of a children’s story comprising 80–120 kana and kanji mixed words. To avoid depending on lexical retrieval to read kanji words, the kanji scripts in the paragraph reading task were accompanied by kana reading (furigana).

##### Picture naming

The patients were asked to name the target picture from six familiar objects. The six items in each category were printed in black on a white card and were shown simultaneously.

##### Spoken word-picture matching

From its spoken name, patients were asked to touch the target picture from six-line drawings (the same as the items used for picture naming).

##### Reading kanji/kana words

The patients were asked to read aloud the target word from six kanji or kana words. Written names of the six familiar items were presented either in kanji or kana scripts (on separate trials). All six words were written in black on a white card and were presented to the patients to read.

##### Spoken verbal command

The patients were asked to make gestures following a simple spoken sentence (e.g., “Open your mouth.”).

### 2.3 Principal component analysis (PCA)

We detected 341 language electrodes in 12 patients. The coverage of language-related electrodes, amongst all electrodes assessed, is shown in Figure 1. Among them, 313 electrodes were assessed with all six tasks and, for clinical reasons, the remaining 28 electrodes were assessed using 4 or 5 tasks.

**Figure 1.**
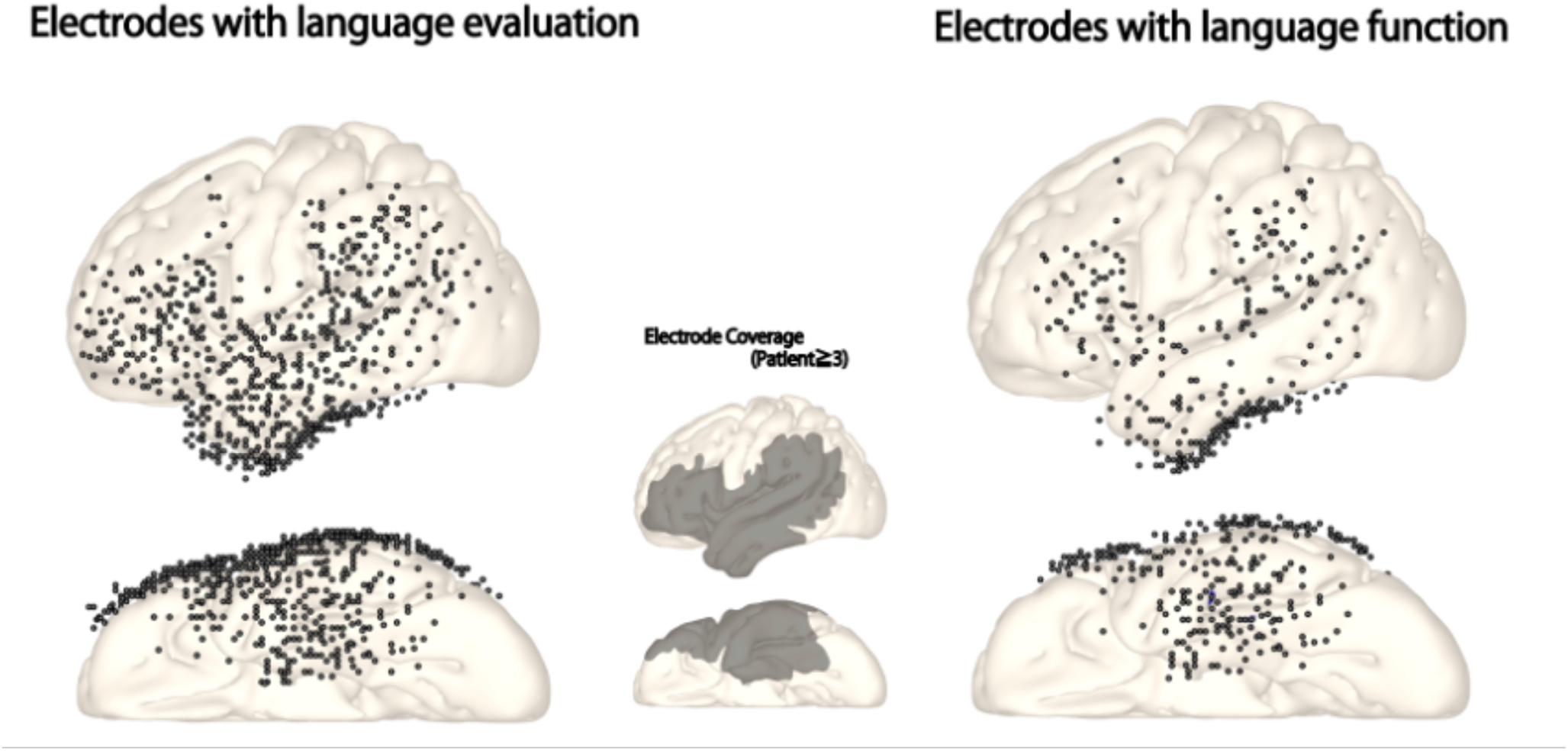
Distribution of the electrode with language evaluation and language function. Fig. 1: Left is the distribution of the electrodes language functions of which are evaluated using 50 Hz stimulation. The middle is the whole electrode coverage. The area where more than 2 patients were included. Right is the distribution of the electrode with language function detected by 50Hz stimulation. Note that the precentral sulcus and postcentral sulcus results are excluded from language mapping because these areas are not precisely evaluated due to their sensory or motor function.

We performed PCA of the 313 language electrodes, which were evaluated using all the six abovementioned language tasks. Participants’ scores (0–2 points) on all language tasks were entered into a PCA by varimax rotation using JMP Pro Statistical Software version 14 (JMP Statistical Discovery LLC, Cary, North Carolina, USA). There are no clear guidelines on the number of cases required for PCA; nevertheless, good results have been obtained using a participant-to-variable ratio of 1:2 (Barrett, 1981). The current study had 6 variables and 313 electrodes, making the ratio 52:1. In addition, factor recovery beyond a sample size of 20 is recommended (Preacher & MacCallum, 2002). Our data, therefore, seem adequate for the purposes of PCA. Factors with an eigenvalue >0.9 were extracted and rotated. After orthogonal rotation, the factor loadings of each test enabled interpretation of what each component represented in the language function evaluated by language tasks.

For the three PCs extracted by the PCA, six tasks were assigned to each PC without duplication based on the factor loading. Next, the PC score was defined as the average of points evaluated in the functional mapping (0–2 points) of the included tasks in each component. For example, if task A and task B are assigned to PC X, the PC score of component X on the specific electrode is (task A’s point + task B’s point) divided by 2.

### 2.4 Region of interest (ROI)

Next, for a clear interpretation of the anatomo-functional relationship of language, we visualized the result of the PC scores on a three-dimensional (3D) brain map and evaluated the result with respect to 10 ROIs. These ROIs were set in each language area to evaluate the functional differences among the subregions of the classical language area. Focusing on the high point area in the language mapping, we referred to anatomical parcellation in the individual brain and obtained the two ROIs in Broca’s area or anterior language area (anterior part of the inferior frontal gyrus [aIFG], posterior part of the inferior frontal gyrus [pIFG]); two ROIs in the anterior part of the lateral temporal area (anterior part of the superior temporal gyrus [aSTG], anterior part of the middle and or inferior temporal gyrus [aMTG/ITG]); three ROIs in the lateral temporo-parietal area, namely, the posterior language area (SMG, posterior part of the superior temporal gyrus [pSTG] and posterior part of the middle temporal gyrus [pMTG]); and three ROIs in the basal temporal area (anterior part of the VTC [aVTC], middle part of the VTC [mVTC], and posterior part of the VTC [pVTC]), for a total of 10 ROIs. In this study, we defined BTLA in a broad sense, which included the entire VTC (Mani et al., 2008). Of note, this definition is different from the classical definition of BTLA suggested by Luders, namely, <6 cm from the temporal tip, and the aVTC and mVTC of our data include the area defined in Luders’ definition of BTLA.

### 2.5 Visualization: comparison between regions

#### 2.5.1 Three-dimensional (3D) map

Obtaining a clear understanding of the whole distribution of language function from the multiple tasks is challenging because electrode locations vary from patient to patient. To understand the spatial distributions of the data derived from all patients, we visualized the data in the Montreal Neurological Institute (MNI) standard space. For each PC, all PC scores were plotted in the MNI space, smoothed using a Gaussian kernel to visualize point data in 3D space. In this study, the 3D volume image that represented the required PC score was averaged across all participants; however, doing so is not simple because electrode location differs by participant. To solve that, we first accumulated the PC score at all electrodes in the MNI space (voxel size, 2 mm; isometric) across all participants, plotting each recorded value (PC score) at the voxel of electrode location. Patient 9 had subdural electrodes embedded in the right hemisphere; therefore, the MNI x-coordinates of this patient were flipped into the left hemisphere. Next, we smoothed the volume (accumulated potential data) using a Gaussian kernel (full width at half maximum, 10 mm; kernel size, 20 mm) to embody the point data in 3D space. In this way, we obtained the volume image of the accumulation of recorded potentials, called the “accumulated score map”. Similarly, we obtained the observation density map by plotting 1 instead of the PC score in each electrode. The accumulated score map was divided by the observation density map to correct for the difference in the electrode density at the individual level. The result is called an “averaged score map”. The calculation was skipped when the observation density was zero. We applied spatial smoothing with a Gaussian filter (full width at half maximum, 10 mm) to the averaged score map. We set the full width at half maximum at 10 mm according to the inter-electrode distance, which is the expected resolution we expect when we perform functional mapping using grid electrodes.

#### 2.5.2 ROI analysis

A 3D map was used to illustrate the PC scores or functional pattern of each language area. Next, we tested the significance of the functional difference between 10 ROIs within 4 language areas to clarify the functional feature of each anatomical region.

A two-way analysis of variance (ANOVA) was performed to test for any significant differences between the combination of PC scores among different ROIs. If the ANOVA was found to be significant, post-hoc tests were conducted to determine the ROIs that had a significantly different pattern of PC score. In addition to comparing ROIs across the whole brain, we investigated significant differences within four areas (the anterior language area, posterior language area, basal temporal language area, and anterior part of the lateral temporal area). The Greenhouse–Geisser correction was applied if sphericity was violated. Unless otherwise stated, a Family Wise Error-corrected p < .05 at the cluster-wise level was applied.

## 3 Results

### 3.1 PCA

The PCA revealed three PCs, explaining 74% of the total variance (Table 2). The KMO (Kaiser-Meyer-Olkin) measure of sampling adequacy was .746, and Bartlett’s test of sphericity was highly significant (p<.001). The factor loadings after rotation are presented in Table 3. Tasks that were related to the reading function loaded heavily on PC1; hence, we refer to this factor as *Reading*. PC2 was interpreted as *Auditory receptive semantic processing*. This is because the tasks that loaded heavily on it were picture–word matching and verbal command (auditory), which were input and interpreted verbally and auditorily, comprehended semantically, and then output non-verbally. The task that loaded most heavily on PC3 was picture naming, followed by kanji word reading; therefore, we assessed PC3 as *Expressive semantic processing*, given that kanji reading has been shown to depend heavily upon semantic-to-phonological output (Fushimi et al., 2009).

**Table 2.** PCA results (Eigenvalue and accumulated contribution rate)

|  | Eigenvalue | Accumulated<br>contribution rate |
| --- | --- | --- |
| PC1 | 2.593 | 43.218 |
| PC2 | 0.954 | 59.114 |
| PC3 | 0.944 | 74.851 |
| PC4 | 0.65 | 85.683 |
| PC5 | 0.454 | 93.245 |
| PC6 | 0.405 | 100 |

**Table 3.**
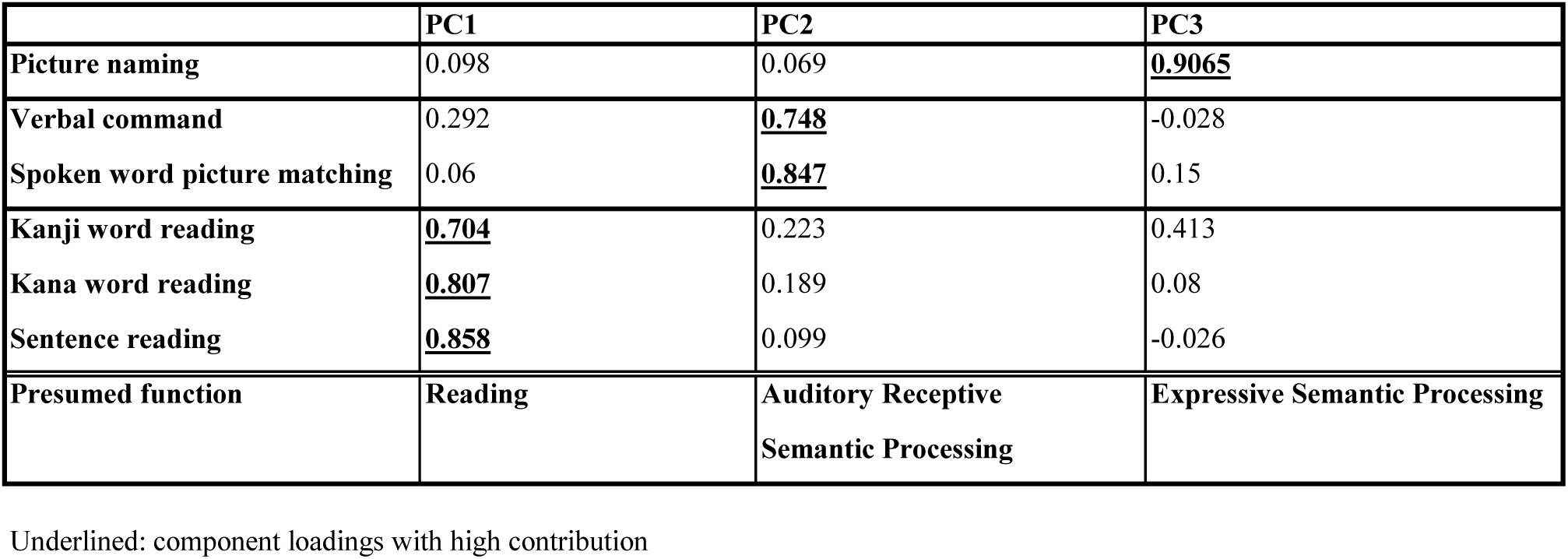
PCA results.

|  | PC1 | PC2 | PC3 |
| --- | --- | --- | --- |
| Picture naming | 0.098 | 0.069 | <u>0.9065</u> |
| Verbal command | 0.292 | <u>0.748</u> | -0.028 |
| Spoken word picture matching | 0.06 | <u>0.847</u> | 0.15 |
| Kanji word reading | <u>0.704</u> | 0.223 | 0.413 |
| Kana word reading | <u>0.807</u> | 0.189 | 0.08 |
| Sentence reading | <u>0.858</u> | 0.099 | -0.026 |
| Presumed function | Reading | Auditory Receptive<br>Semantic Processing | Expressive Semantic Processing |

Underlined: component loadings with high contribution

Table 3: Loading from PCA. PC1 is related to Kanji, Kana and Sentence reading. PC2 shows a high value for verbal command and spoken word picture matching. PC3 is highly related to picture naming.

### 3.2 ROI analysis

Figure 2 shows the averaged PC score map and each ROI. The PC1 score was prominent in the posterior part of the basal temporal lobe and pars opercularis (pOpe). A high PC2 score was observed in the posterior and superior parts of the lateral temporal lobe and the anterior part of the basal temporal lobe. Compared to PC2, SMG showed a relatively high PC1 score; the PC2 score in the pars triangularis (pTri) was higher than the PC1 score. Unlike the PC1 and PC2 scores, the PC3 score was high over a broad area, mainly the base of the temporal lobe along the occipitotemporal sulcus, and the SMG, pMTG, and pOpe. The sum of the PC1, PC2, and PC3 scores showed a similar distribution to the PC3 score or picture naming task score.

**Figure 2.**
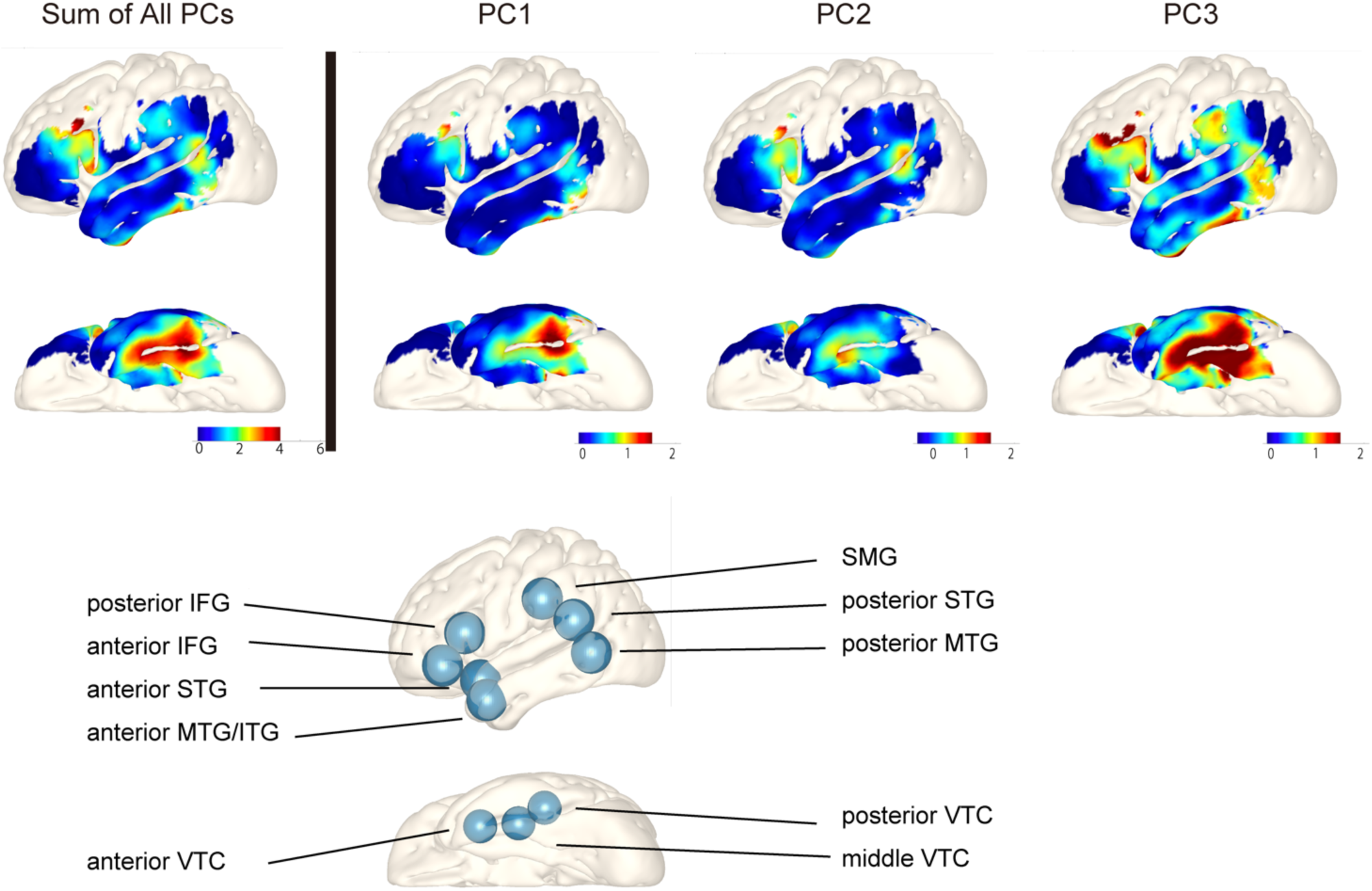
Results of PC scores visualized on the cortex and regions of interest. Fig. 2: The upper row is the visualization of each PC’s score and the sum of all PC’s scores on the brain image. The colour scale is shown under each brain map. PC1 is prominent at the posterior part of the ventral temporal cortex (VTC) and pars opercularis, PC2 at the posterior part of the superior temporal gyrus (STG) and the anterior part of VTC, and PC3 at the supramarginal gyrus (SMG), the posterior part of middle temporal gyrus (MTG) and whole VTC. The lower row shows 10 regions of interest, 7 from the lateral surface of the frontal, temporal and parietal lobes, and 3 from the base of the temporal lobe.

Two-way repeated ANOVA was conducted for each PC and all 10 ROIs in the language areas. The analysis revealed a significant difference among 10 ROIs (Figure 3A). Next, ANOVA for ROIs within each language area (including the anterior part of the lateral temporal area, and the anterior, posterior, and basal temporal language areas) and PCs (including PC1, PC2, and PC3) was conducted (Figure 3B). This second analysis showed significant interaction among three ROIs within the posterior language area and the BTLA. This result suggests that both the posterior and basal temporal language areas have functionally segregated subregions within each area. Post-hoc analysis revealed significantly different patterns of PC scores in the following pairs: between SMG and pSTG in the posterior language area, and between aVTC and pVTC, as well as between mVTC and pVTC, in the basal temporal area.

**Figure 3.**
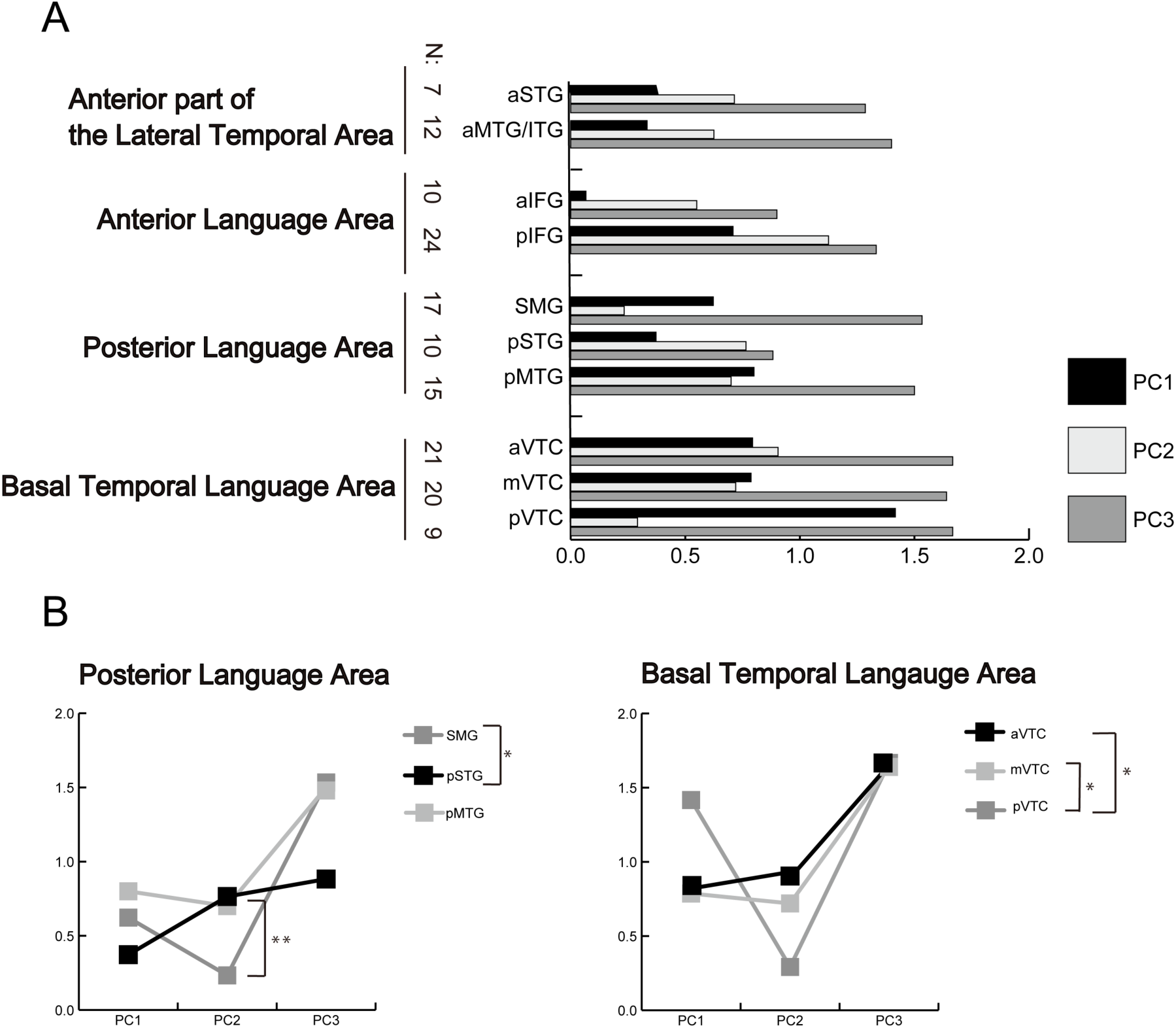
Results of ANOVA on 10 ROIs, posterior language area, and basal temporal language area. Fig. 3: The upper graph(A) shows the result of two-way repeated analysis of variance (ANOVA) on the PC scores and 10 ROIs. ANOVA showed significant interaction between PCs and ROIs. Post hoc analysis (B, the lower 2 graphs) revealed functional differentiation within the posterior language area (low PC1 at the posterior STG, and high PC3 at the posterior MTG and SMG) and BTLA (high PC1 and low PC2 at the posterior part).

In the posterior language area, the comparison between SMG and pSTG revealed a contrasting pattern for PC1 and PC2 (PC1>PC2 in the SMG and vice versa in the pSTG) and lower PC3 score in the pSTG than in the SMG. With regards to the pMTG, it tended to have higher PC1 and PC3 scores than the adjacent pSTG. In the BTLA, pVTC was characterized by a higher PC1 and lower PC2 score pattern. A remarkably high PC3 score was observed in all BTLA subregions.

The second ANOVA analysis did not reveal any significant interaction among regions for the anterior language area and the anterior part of the lateral temporal area. In the anterior language area, the patterns of the three PC scores were similar (PC1<PC2<PC3) between the anterior and posterior parts of the IFG; however, PC1 was smaller in the aIFG, and the pIFG had high scores as a whole. In the anterior part of the lateral temporal area, the patterns of the three PC scores were similar in both ROIs, namely, the aSTG and aMTG/ITG.

## 4 Discussion

### 4.1 Summary

The salient results of the study were as follows: 1) Based on the language function mapping results obtained during electrical cortical stimulation, we extracted three principal component (PCs) variations from multiple language tasks. 2) We visualized each PC score and identified statistically significant differences in the distribution patterns of each function across language-related regions. 3) Significant differences were observed in functional patterns between subregions, especially in the posterior language area and BTLA. This included differences in the functions of the pSTG, pMTG and SMG within the posterior language area, as well as differences in the functions of the anterior and posterior parts of the VTC.

### 4.2 Anatomo-functional properties of each PC

In the present study, three PCs were extracted. The function represented by each component was interpreted while considering the phonological and semantic aspects and nature of each language task. Akin to this study, employing PCA, Halai et al. utilized data from patients with post-stroke aphasia to extract components of speech fluency, phonology, and semantics from neuropsychological task performance, further correlating them with lesion distribution (Halai et al., 2017). Similarly, our approach successfully drew from the direct performance outcomes of electrical cortical stimulation, extracting each language function from semantic and phonological perspectives. PC1, associated with reading sentences and kanji and kana words, was interpreted as supporting reading. PC2, related to verbal commands and word–picture mapping, was understood to support auditory processing of receptive semantic tasks. PC3, which involved naming, with some indications pointing toward some involvement with kanji reading, was considered to support expressive semantic processing. Due to the nature of PCA, the interpretation of PCs is operational; however, if the assumed functions are consistent with the cortical distribution, the interpretation of the PCs is supported.

PC1 showed high loadings for kanji and kana word and sentence reading. Components of the reading function, which involve phonology, vision and speech, was extracted on PC1. The unique aspects of the Japanese language govern an individual’s recognition of kana letters as phonetic characters and kanji letters as ideographic characters. It should be noted that in the tasks used in this study, kanji stimuli were limited to the characters representing consistently-pronounced words that do not always require semantic processing and have only one pronunciation(Fushimi et al., 2009). Additionally, sentence reading included kanji characters that were accompanied by kana characters (furigana) indicating how to pronounce each word; thus, lexical-semantic retrieval was not required to read these kanji words (Matoba et al., 2024). Therefore, the scores on PC1, probably reflects the elements of reading related to phonology, vision and speech, rather than semantic processing.

In the present study, reading networks were found in the pVTC, pMTG, pOpe, and SMG. The pVTC includes a visual word form area (vWFA) located in the posterior part of the fusiform gyrus, and the pMTG has been proposed as an interface between the phonological and semantic systems (Binder, 2017). A distribution consistent with a pathway from visual recognition to the integration of phonological and semantic information is relevant for reading comprehension (Chen et al., 2019; Cohen et al., 2000; Cohen et al., 2008; Hickok & Poeppel, 2007; Jobard et al., 2003). Numerous studies have shown that the vWFA, a part of the posterior VTC, is involved in the first step of reading processing, i.e., the recognition of visual information about word forms (Cohen et al., 2002; Hodgson et al., 2021; Price & Devlin, 2003; Reinholz & Pollmann, 2005; Turkeltaub et al., 2002; Vigneau et al., 2005). Multiple findings also suggest that the lexico-semantic processing route, which converts spelling to phoneme and semantics, corresponds anatomically to the pathway from the pITG and pMTG to the aIFG. Similarly, the phonological decoding process spans anatomically from the left STG, ventral IPL, and SMG and extends to the precentral gyrus and pIFG (Price, 2012). The distribution for the left ITG, pMTG, SMG, and IFG observed in PC1 aligns with the established reading network.

PC2 was strongly associated with verbal commands and word–picture matching tasks. Both involve auditory verbal input, comprehension of meaning, and transformation into a non-verbal output; thus, PC2 is indicative of the function of auditory processing of receptive semantic tasks. Therefore, it suggests the involvement of regions related to auditory comprehension. PC2 is strongly associated with the pOpe, pTri (both located in the left IFG), pSTG, and aVTC. These regions play an important role in auditory semantic comprehension, as noted in previous studies (Mani et al., 2008; Vigneau et al., 2006). The pSTG is particularly important for the phonological analysis of auditory input and is responsible for the extraction of phonological features in the early stage of comprehension (Robson et al., 2012). The anterior part of the VTC, or ventral ATL in many studies, also plays a central role in the integration of semantic information, serving as a hub for conceptual information of multiple modalities (Ralph et al., 2017; Shimotake et al., 2015). Impairment in PC2-related tasks manifests with symptoms that closely resemble Wernicke aphasia or pure word deafness. Indeed, the region associated with the PC2 distribution closely matches the region of damage (pSTG and pMTG) in Wernicke aphasia (Robson et al., 2012). Thus, PC2 distribution is assumed to function together to form a network that supports auditory semantic comprehension.

PC3 categorized naming with extremely high factor loading and showed a slightly high association with kanji reading tasks. Statistically speaking, PC3 represents the residual (orthogonal) variance in naming after accounting for variations on PC1 and PC2. Hence, PC3 is seems to relate to additional variation in expressive semantic processing. The pMTG-ITG, SMG, IFG, and the whole VTC were the sites where the PC3 map showed association. The pMTG is important for flexible, executively-related processing of semantic information which is required for all demanding semantic tasks including semantic speech production and naming (“semantic control”: (Hodgson et al., 2021; Jefferies & Lambon Ralph, 2006; Noonan et al., 2013). Damage to the pMTG and pITG lead to impairments of semantic control in the posterior subgroup of semantic aphasia and Wernicke’s aphasia (Thompson et al., 2015). SMG may be associated with speech in the naming task due to its function in phonological processing (also compatible with a lower association in PC2 without speech). The left IFG seems to house both the anterior regions (that work in concert with posterior lateral areas) for semantic control (ref(Hodgson et al., 2023; Jefferies & Lambon Ralph, 2006; Noonan et al., 2013)), and phonological processing in neighbouring subregions (Shekari & Nozari, 2023). The BTLA (or VTC in this study) is important for visual recognition and semantic processing as shown in neurostimulation studies (Cohen et al., 2000; Hoffman et al., 2015; Shimotake et al., 2015) as well as the anterior VTC being the peak of atrophy in semantic dementia and known to be a centre-point for multimodal semantic representations (Ralph et al., 2017). Overall, the results of the current study are in good agreement with those of classical naming stimulation studies, with respect to the frontal and temporal lobes (Ojemann et al., 1989) and basal temporal lobe (Luders et al., 1991). Overall, the network of brain regions implicated here with semantic representation, naming and semantic control mirror those found in a recent large meta-analysis of the functional neuroimaging literature (Hodgson et al., 2023).

### 4.3 Functional features of each language area as evaluated using PCs

Evaluating each language area with the PC profile revealed distinct features for each area and functional differentiation within the regions of the language areas. No interaction was found between two ROIs of the anterior language area (i.e., the IFG) after conducting the ANOVA. Both the aIFG and pIFG followed the pattern PC1<PC2<PC3, but the overall strength of language function was significantly greater in the pIFG than in the aIFG. The left pOpe and pTri are regions that support auditory working memory (Burton et al., 2000; Hsieh et al., 2001). The pIFG, or BA44, is considered to be related to articulatory aspect of speech processing (Binder et al., 2004; Burton et al., 2005; Moser et al., 2009). This region is suggested to have a strong association with phonological function: the high PC1 scores observed in this region in the current study corroborate this notion. Our study also revealed the probable semantic function of the pTri, assumed by the difference in PC1 and PC2 scores. Indeed the BA44—and at times the BA45 (pIFG in the present study)—is involved in semantic processing (Hoffman et al., 2010; Jefferies & Lambon Ralph, 2006; Nakai et al., 1999) or in lexical decision-making (Perani et al., 1999). Semantic processing or retrieval has been associated with the anterior part of the IFG (pars orbitalis and pTri). Conversely, in our study, the pars orbitalis was not related to any function among the three PCs. A possible explanation for this discrepancy is that the semantic tasks set in this study were very simple and did not require a high level of semantic function.

Different functional patterns were found between the three ROI subregions of the posterior language area. The SMG and pMTG exhibited the pattern PC3>1>2, and the pSTG exhibited the pattern PC3≥2>1 (Figure 3). Multiple comparisons for each PC revealed a significant difference between the PC2 scores for the pSTG and SMG, with the low values of the SMG being noteworthy. Specifically, the SMG was strongly associated with PC3 (expressive semantic processing or naming function), while its involvement in PC2 (auditory comprehension and receptive semantic processing) was small. The pSTG was equally involved in semantic processing for both PC2 and PC3. pMTG was associated with both PC1 (phonology) and PC2 and PC3 (both semantic) functions.

Functional imaging studies have pointed to the integration of semantic and phonological information as a function of the pSTG (Hodgson et al., 2023), and Binder et al. posited that the pSTG is involved in the maintenance of phonological representations and with phonological short-term memory tasks(Binder, 2017). The ROI of the pSTG in this study is located posterior to the region marked by the classic narrow Wernicke’s area (Tremblay & Dick, 2016). Although it is agreed that Wernicke’s area is involved in phonological processing and auditory short-term memory (Buchsbaum et al., 2001; Leff et al., 2009; Richardson et al., 2011), its anatomy and function remain controversial (Binder, 2017). A prior study using voxel-based lesion-symptom mapping revealed that auditory comprehension deficits were not associated with the posterior portion of BA22, traditionally known as Wernicke’s area. Instead, lesions in a more posterior region, specifically in the posterior STG and MTG, surrounding the posterior end of the superior temporal sulcus were significant for auditory comprehension (Dronkers et al., 2004). Indeed, by comparing Wernicke’s and transcortical sensory aphasia (semantic aphasia) directly, Robson et al. found that the comprehension impairment shared by both patient groups was associated with the pMTG whilst the extension to pSTG and inferieor SMG in the WA patients was associated with their poor auditory-phonological and working memory deficits (Robson et al., 2012; Thompson et al., 2015). Our findings provide electrophysiological support for these observations and suggest that the very posterior section of the pSTG could be considered a more precisely defined Wernicke’s area, as a region of receptive phonology.

The association of the SMG with semantic expression or naming is also consistent with electrical cortical stimulation studies (Ojemann et al., 1989). Accumulated fMRI data have also indicated activation of the SMG in the naming function (Oberhuber et al., 2016). Lesion studies also showed involvement of the SMG during object naming (Fridriksson et al., 2018), especially in phonological processes (Dell et al., 2013). Oberhuber et al. have also shown that multiple subregions of the SMG are involved in the reading and naming functions (Oberhuber et al., 2016), consistent with the PC1 and PC3 maps (Figure 2). The reason for the lack of association of the SMG with PC2, despite this region being implicated in both semantic and phonological processing, may be that the PC2-related tasks (verbal command and word–picture matching) did not require verbal responses, or that the tasks were easier (compared to semantic tasks in general).

The present PCA findings revealed that while the pMTG partially contributes to phonological processing, it primarily plays a role in semantic system. As previously noted, the pMTG has long been considered to be involved in auditory comprehension and phonological processing streams (Bates et al., 2003; Dronkers et al., 2004; Hickok & Poeppel, 2007; Price, 2012). On the other hand, recent studies have demonstrated that the pMTG plays a central part in the semantic network, particularly as a key region for semantic control, in contrast to the multi demand network (Davey et al., 2016; Jackson, 2021; Noonan et al., 2013). Although this study was not specifically designed to evaluate semantic control, the prominence of the pMTG in both PC2 and PC3 is consistent with these findings.

The present study showed that language functions within the BTLA also differed by subregion (anterior, mid, and posterior). Naming or expressive semantic processing was strongly associated with all parts of the BTLA, which is thought to play an important role in language processing by electrical cortical stimulation (Luders et al., 1991). In particular, electrical stimulation of the BTLA elicits generalized receptive and expressive aphasia. A previous study indicated that the entire BTLA is involved in the integration of visual and auditory information, semantic processing, and phonological processing (Chen et al., 2016; Krauss et al., 1996; Ralph et al., 2017), and recent studies have also shown amodal characteristics of the anterior half (Matoba et al., 2024), but few in-depth studies exist on the functional differences between the anterior and posterior parts of this area directly. Considering that the posterior region encompasses the vWFA and is part of the visual ventral stream (called the “*what pathway”*), it is plausible that this region showed a high level of impairment in tasks requiring text reading and object naming. The anterior VTC is the most prominent area of atrophy in semantic dementia, which is associated with semantic deficits in multimodal expressive and receptive semantic deficits. Functional imaging, rTMS, and ECS studies have shown the same findings, and anterior VTC is suggested as the semantic region. (Lambon Ralph et al., 2010; Mion et al., 2010; Patterson et al., 2007; Ralph et al., 2017).

Due to the small sample size, no statistically significant results were obtained in the anterior part of the lateral temporal area (i.e., the aSTG and aMTG/ITG), nor were visual differences in functional patterns observed between the subregions on PC maps (Figure 2). PC1 showed lower scores and relatively stronger associations with semantic processing, especially in comparison with PC3. This is consistent with previous findings of ATL involvement in name mapping (Damasio et al., 2004) and does not contradict the role of the ATL as a semantic hub.

### 4.4 Clinical implications: task selection

This study has clinical significance as, using PCA, we were able to achieve dimension reduction of the mapping results obtained from six tasks into three groups. From the results of the PCA, selecting one task from each PC group (i.e. PC1, PC2, and PC3), for a total of three tasks (e.g. sentence reading from PC1, picture matching from PC2, and naming from PC3), could substitute the results of all six tasks. This is considered a useful outcome for achieving minimally invasive mapping. This ingenuity of electrical cortical stimulation (ECS) mapping can be applied not only to the presurgical evaluation with subdural electrodes but also to that with stereo-EEG and can improve efficiency. Task reduction is also useful clinically for efficient language mapping during the limited time frame of awake craniotomy for brain tumor surgery.

### 4.5 Limitations of the study

The first limitation of this study was the restricted range of electrode coverage. The evaluation was limited to the cortex where electrodes were placed. By selecting cases that included known language areas and where comprehensive stimulation evaluation was performed on all electrodes, the study aimed to ensure as much reliability as possible.

Second, the absence of a repetition task must be noted. The six tasks used in this study did not include any in which semantic processing was completely bypassed though kana reading is fairly close; thus, the study lacked a pure assessment of phonology. While the inclusion of semantic processing is indispensable for evaluating the localization of comprehensive language functions, and therefore its omission from this study is rational, the addition of tasks such as non-word repetition limited to the posterior language area may be considered in clinical task selection.

## 5 Conclusion

The ECS mapping results of the six language tasks could be grouped into three PCs, which showed valid localizations supported by PCA. By evaluating the PC profile of each language area, the functional differences between language-related regions, especially the functional subdivision of the posterior language area, could be directly demonstrated. Clinically, task selection, considering PCA, allows for mapping with less evaluation time and zero loss of information compared to mapping using more tasks, and yields beneficial results.

## Data availability statement

All data used for the analyses reported in this manuscript are available upon request.

## Author Contributions

Mayumi Otani (Conceptualization, Data curation, Formal Analysis, Investigation, Methodology, Project administration, Resources, Software, Visualization, Writing—original draft, Writing—review & editing), Akihiro Shimotake (Conceptualization, Data curation, Formal Analysis, Funding acquisition, Investigation, Methodology, Project administration, Resources, Supervision, Writing—review & editing), Takuro Nakae (Conceptualization, Formal Analysis, Investigation, Methodology, Project administration, Software, Supervision, Visualization), Mitsuhiro Sakamoto(Data curation, Formal Analysis, Investigation, Methodology, Supervision), Masao Matsuhashi (Conceptualization, Data curation, Formal Analysis, Funding acquisition, Investigation, Methodology, Project administration, Resources, Software, Supervision, Visualization), Takayuki Kikuchi(Data curation, Funding acquisition, Methodology, Resources, Supervision), Kazumichi Yoshida (Data curation, Resources, Supervision), Takeharu Kunieda (Conceptualization, Data curation, Funding acquisition, Methodology, Project administration, Resources, Supervision), Susumu Miyamoto (Conceptualization, Data curation, Investigation, Methodology, Project administration, Resources, Supervision), Ryosuke Takahashi (Project administration, Resources, Supervision), Matthew A Lambon-Ralph(Conceptualization, Formal Analysis, Methodology, Project administration, Supervision, Writing—review & editing), Akio Ikeda (Conceptualization, Data curation, Funding acquisition, Investigation, Project administration, Resources, Supervision, Writing—review & editing), and Riki Matsumoto (Conceptualization, Data curation, Formal Analysis, Funding acquisition, Investigation, Methodology, Project administration, Resources, Supervision, Writing—review & editing).

## Acknowledgements

We gratefully acknowledge the other neurologists and neurosurgeons at Kyoto University who contributed to the presurgical evaluations.

